# Rapid Detection of *Candida auris* Using a Loop-Mediated Isothermal Amplification (LAMP) method

**DOI:** 10.1101/2025.03.04.25323353

**Authors:** Leila Masoumi, Hamed Fakhim, Mohammad Pourhassan-Moghaddam

## Abstract

**Background:** *Candida auris* (*C. auris*) can cause nosocomial transmission in susceptible hosts. This study aimed to use the Loop-Mediated Isothermal Amplification (LAMP) technique as a novel detection technique for *C. auris* and compare the efficacy with PCR

**Methods:** This study was performed on *C. auris* fungal isolates as positive samples and five different standard *Candida* species as negative control samples. The sensitivity and specificity were determined using different *C. auris* DNA concentrations. LAMP products were identified by electrophoresis, turbidity, and colorimetric methods. Finally, the products were sequenced

**Results:** The LOD (limit of detection) of the LAMP was 10 times lower than the PCR reaction. LAMP and PCR correctly identified *C. auris* with 100% specificity. The nBLAST results showed the similarity between the sequences of the PCR products and Gene Bank sequences, confirming the specific amplification of target gene in *C. auris* genome by LAMP and PCR methods.

**Conclusion:** The reproducibility and reliability of the LAMP reaction in differentiating *C. auris* from closely related species were confirmed. The developed LAMP technique holds promise for testing the clinical samples for quick detection of *C. auris* with excellent accuracy.

## 1. Introduction

*Candida* genus is the most common fungal pathogen causing serious healthcare-associated infections such as Candidemia in the bloodstream, especially in patients admitted to the ICU (intensive care unit) [1]. *Candida auris* (*C. auris*) has been considered one of the most important species of invasive Candidal infections since 2015. It is defined that *C. auris* is multi-drug resistant and has a high rate of spread and mortality rate [2]. Despite the development of PCR (polymerase chain reaction), a powerful method for the diagnosis of infectious diseases, it takes more time to detect which increases inappropriate antimicrobial therapy and mortality rate induced by infections. In addition, detection of some fungal species cannot be performed by *in vitro* culture methods. Therefore, there is a need for point-of-care testing (POCT) which is a faster, more sensitive, and specific diagnostic method for infectious diseases instead of cell culture-based methods [3–6].

Nucleic acid testing techniques directly trace DNA sequences, allowing more clinical information to be gleaned from the patient or pathogen. Antimicrobial resistance, biomarkers of virulence, and type of pathogen can be quickly identified so that optimal treatment and therapeutic intervention can continue without any delay. Also, the quantitative evaluation of the pathogen load has a strong prognostic value [7]. The advent of PCR revolutionized genetics and molecular diagnostics by providing a simple and elegant method for nucleic acid amplification using heat-resistant polymerase enzymes and alternating temperature cycles for strand separation and amplification. This powerful technology is widely used for molecular diagnostics, biomedical research, and life sciences [8]. Unfortunately, the precise and repetitive heating cycles required for PCR necessitate a complex design and make PCR an incomplete solution for integration into POCT systems. Confining a PCR reaction in an enclosed microfluidic system requires additional engineering considerations that are limited by the thermal constants of the materials used, the heat sensitivity of many enzymes used in detection, unwanted water evaporation, and complex thermal control mechanisms. In order to circumvent the limitations of traditional PCR in the molecular diagnostic amplification steps of POCT, recent research has been directed towards isothermal methods for nucleic acid amplification [9]. Isothermal amplification techniques use enzymes to perform strand separations that would otherwise require repeated heating to achieve. These techniques offer those developing POCT detection platforms a powerful tool for the amplification and detection of nucleic acids without the traditional complexity of thermally repetitive steps and associated control mechanisms. Thus, one of the most sensitive types of isothermal methods, the LAMP method, was used in this study to identify *C. auris*.

Since the isothermal amplification technique using the modern molecular method ring is highly sensitive in detecting infectious agents and low cost (no need for thermocycler and electrophoresis), the LAMP technique could be a proper replacement for conventional diagnosis methods. So, this study proposed investigating if LAMP would differentiate *C. auris* from other *Candida* species. Moreover, the results obtained were compared with PCR as the molecular gold standard method.

## 2. Materials & methods

### 2.1. Cell culture and DNA extraction

*Candida* fungal isolates were cultured on Sabouraud dextrose agar (SDA) medium - containing Peptone, glucose, and agar - at 37 °C for 2 days. Then, the fungal colony was dissolved in 100 µl lysis buffer for cell lysis. Then the samples were boiled at 100 °C for 15 minutes, added potassium acetate, and incubated on ice for 60 minutes. Finally, the samples were centrifuged, the pellet was dried at room temperature for 20 minutes, and stored at -20 °C with 50 µL TE buffer for further analysis.

The quality and quantity of DNA samples were examined via the nanodrop device by measuring optical density (OD) ratio of 260/280 and 260/230.

### 2.2. DNA amplification and confirmation by PCR

In this study, external F3 (5’-ACCCAACGTTAAGTTCAACT-3’) and B3 (5’-CGCGAAGATTGGTGAGAAG-3’) primers designed for LAMP reaction (Macrogen, Korea), were also used for the amplification of the ITS fragment using PCR. The PCR reaction containing F3 and B3 primers and other components (PCR maxtermix, Cinnagen, Iran) were run using thermal cyclers (PEQLAB, Germany). The PCR program consisted of denaturation at 94 °C for 5 minutes, and then 30 cycles of denaturation (at 94 °C for 30 seconds), annealing (at 56 °C for 30 seconds), and extension (at 72 °C for 30 seconds) steps. DNA samples were incubated at 72 °C for 10 minutes for final extension.

Finally, 3 µL of the PCR product was electrophoresed in a 1% agarose gel (Cinnagen, Iran) to confirm the 180 bp fragment amplicons and used as the definite positive sample in LAMP.

### 2.3. LAMP

LAMP primers for amplification of ITS-2 region of *C. auris* genome were designed using Primer Explorer V online software (http://primerexplorer.jp/). Primer sequences F3, B3, F1P, B1P, F2, F1C, B2, and B1C were 5’-ACCCAACGTTAAGTTCAACT-3’, 5’-CGCGAAGATTGGTGAGAAG-3’, 5’-GCATTTCGCTGCGTTCTTCATCACATAAAACTTTCAACAACGGAT-3’, 5’-ACTTGCAGACGTGAATCATCGAATACATCACGCTCAAACAGG-3’, 5’-ACATAAAACTTTCAACAACGGAT-3’, 5’-GCATTTCGCTGCGTTCTTCATC-3’, 5’-ACATCACGCTCAAACAGG-3’, and 5’-ACTTGCAGACGTGAATCATCGAAT-3’, respectively.

To check the test’s specificity, the LAMP reaction was performed simultaneously on the extracted genome of *C. auris* (definitive positive control) and 5 negative controls of *Candida* samples.

The LAMP reaction was performed on eight types of extracted *Candida* DNA samples using the Bst DNA polymerase enzyme in a final volume of 25 μL at a temperature of 65 °C for 60 minutes in a heat block (Table 1).

**Table 1.** LAMP reaction ingredients.

| **Materials** | **Value** | **Concentration** |
| --- | --- | --- |
| **Genomic DNA** | 1 µl | 4 ng/µl |
| **LAMP buffer** | 2.5 µl | 1 x |
| **MgSO<sub>4</sub>** | 2 µl | 8 mM |
| **dNTP** | 1.4 µl | 10 mM |
| **Betaine** | 4 µl | 0.8 M |
| **FIP primer** | 2 µl | 40 pM |
| **BIP primer** | 2 µl | 40 pM |
| **F3 primer** | 0.25 µl | 5 pM |
| **B3 primer** | 0.25 µl | 5 pM |
| **Bst DNA Polymerase** | 1 µl | 8 U |
| **H<sub>2</sub>O** | 6.5 µl | - |
| **Total** | 25 µl | - |

LAMP: Loop-Mediated Isothermal Amplification.

### 2.4. The specificity and sensitivity of the LAMP and PCR tests

To determine the specificity of the test, the *Candida* family, including *C. albicans*, *C. glabrata*, *C. krusei*, *C. paraglabrata,* and *C. tropicalis* were simultaneously tested with positive samples in the LAMP reaction as negative control samples.

At last, 3 µL of the product were loaded in agarose gel 1.5% for electrophoresis at 85 V for 60 minutes. The following formula was used to calculate the number of *C. auris* copies:

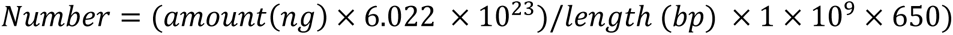

Accordingly, dilutions of 10^3^-10^6^ copies/tube with a dilution factor of 10^-1^, 1, 5, 10, 100, 250, and 500 copies/tube were prepared.

To determine the sensitivity of the reaction, master mix was added to the 1 µL of the amplified DNA samples and incubated in the Thermo Block for 60 minutes at 65°C.

The same dilutions prepared for the LAMP reaction were used to determine the sensitivity of the PCR reaction and the results were analyzed by electrophoresis on a 1% gel.

### 2.5. Observational detection

#### Turbidity

After completing the LAMP reaction, without performing electrophoresis and immediately after the completion of the reaction, the positive sample was compared with the negative sample (*Candida albicans*) in terms of turbidity.

#### Colorimetric detection

For colorimetric detection of samples, 1 µL of hydroxynaphthol blue (HNB) 0.1mM was added at the beginning of the reaction along with the rest of the reaction mixture.

### 2.6. DNA Sequencing and nBLAST

As we used F3 and B3 primers of the LAMP reaction in PCR experiments, the sequencing was only performed on PCR products, which represent same target gene region amplified in both PCR and LAMP reaction. Briefly, DNA sequences obtained from PCR reactions were automatically sequenced using the Sanger method (Applied bioSystems (ABI)). The reading of the sequences was done using Chromas software and reported as a graphic file called chromatogram and a text file.

The sequences similarity was searched in the NCBI database (https://blast.ncbi.nlm.nih.gov) using nucleotide BLAST (nBLAST) tool. nBLAST alignment results reported with criteria (total query coverage, max identity, E value, max score, and total score).

### 2.7. Phylogenetic tree and evolutionary path

The phylogeny tree was created with MEGA 6 software based on the neighbor-joining method, with a bootstrap value of 1000.

## 3. Results

### 3.1. C. auris culture and DNA extraction

The standard candidate strains were cultured in SDA culture medium and incubated for 48 hours. Then, to optimize the LAMP and PCR techniques, DNA samples extracted from them were used as negative and positive control samples during this research.

The OD ratio of 260/280 samples was between 1.8-2. The mean concentration of DNA samples in *C. albicans*, and *C. glabrata*, and *C. krusei*, and *C. parapsilosis*, and *C. tropicalis*, and *C. auris isolate UMS44* (sample 1), *C. auris isolate UMS48* (sample 2), and *C. auris isolate UMS51* (sample 3) were 174.8, 198.4, 101.8, 195.2, 157.7, 167.3, 181.6, and 138.2 ng/µL, respectively. After calculating, the DNA concentration of all samples was diluted to 4 ng/µl.

### 3.2. The specificity of C. auris detection

Amplification and identification of ITS gene of *C. auris* mushroom was evaluated simultaneously with negative control samples by PCR technique and specially designed primers. PCR results of the ITS gene confirmed the presence of a fragment of about 180 bp, which corresponds to a part of the ITS gene, in the electrophoresis gel (Figure 1).

**Figure 1.**
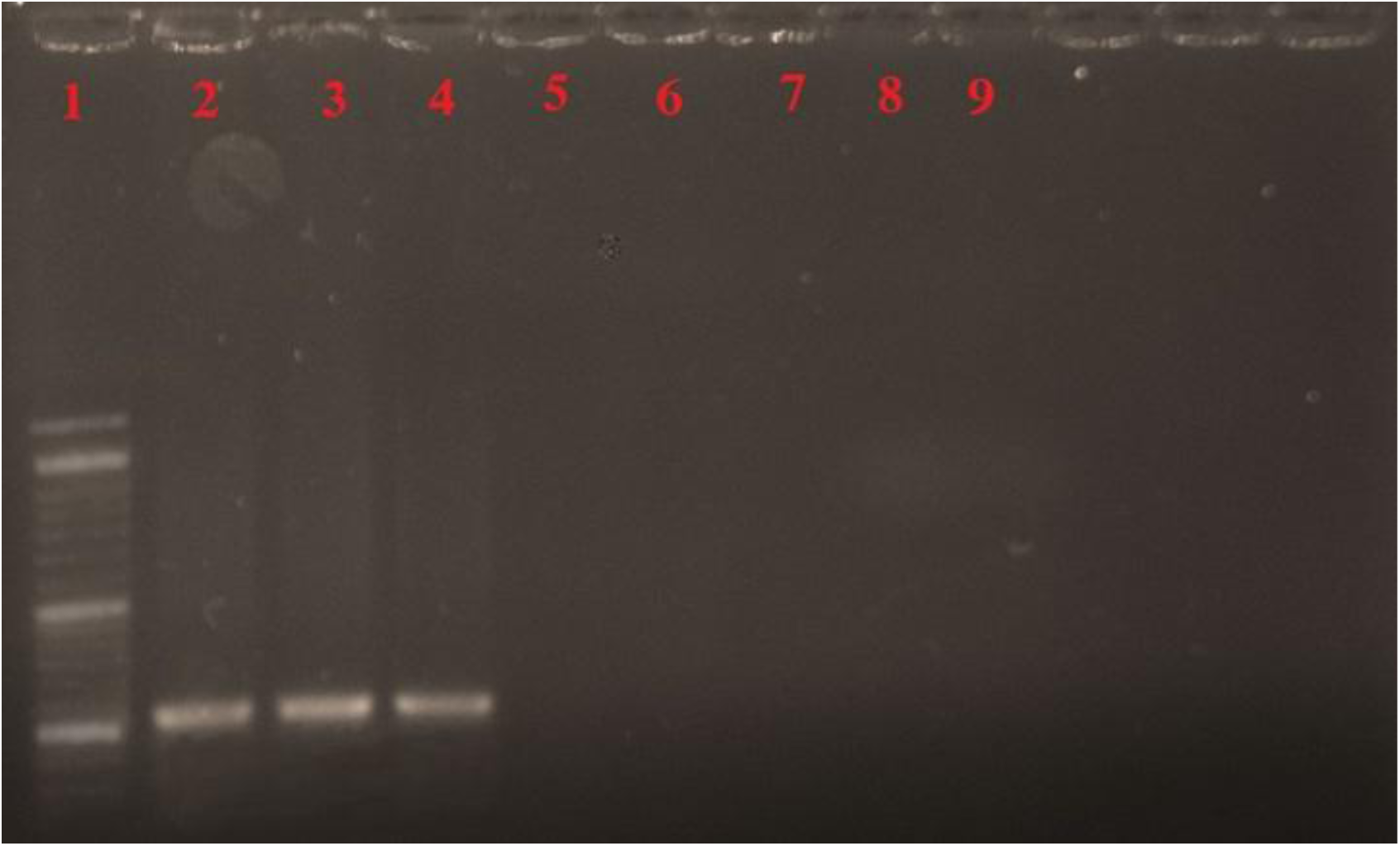
The specificity of *C. auris* detection via PCR method. The gels have 1) Ladder 50-kbp, 2) *C. auris* sample 1 (UMS44), 3) *C. auris* sample 2 (UMS48), 4) *C. auris* sample 3 (UMS51), 5) *C. albicans*, 6) *C. glabrata*, 7) *C. krusei*, 8) *C. parapsilosis*, and 9) *C. tropicalis* samples.

To check the specificity of the LAMP method, DNA samples extracted from *Candida* family including: *C. auris sample 1 (UMS44), C. auris sample 2 (UMS48), C. auris sample 3 (UMS51)* (all three as positive controls), *C. albicans, C. glabrata, C. krusei, C. parapsilosis, and C. tropicalis* (all five as negative controls) were used in LAMP reactions. The results of this study showed that the LAMP reaction was positive only in relation to *C. auris* isolates, which indicates 100% specificity of the reaction at the molecular level (Figure 2).

**Figure 2.**
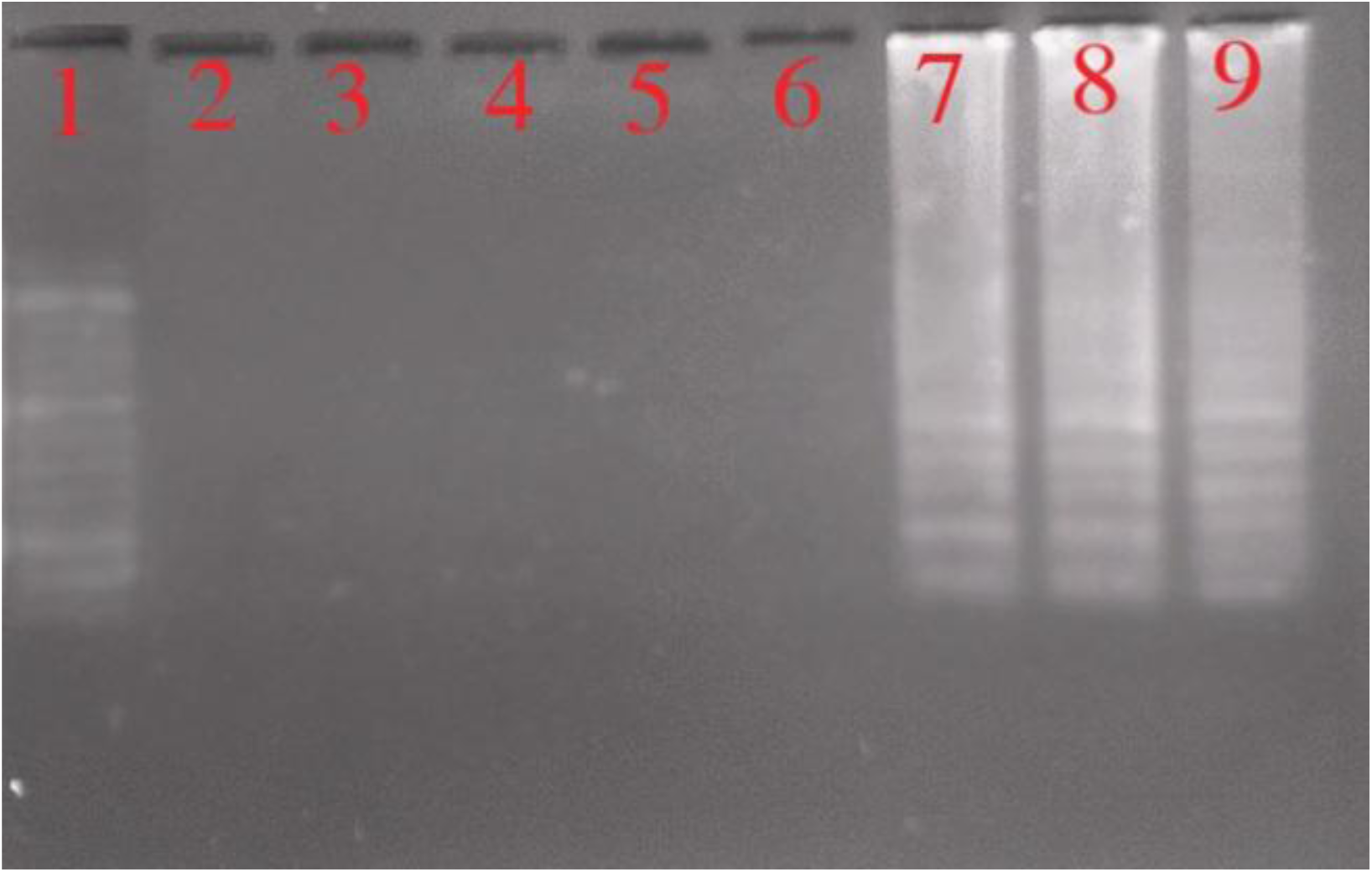
The specificity of *C. auris* detection via LAMP method. The shown products are the 1) Ladder 50-kbp, 2) *C. albicans*, 3) *C. glabrata*, 4) *C. krusei*, 5) *C. parapsilosis*, 6) *C. tropicalis*, 7) *C. auris* sample 1 (UMS44), 8) *C. auris* sample 2 (UMS48), and 9) *C. auris* sample 3 (UMS51) samples.

### 3.3. The sensitivity of C. auris detection

To check the analytical sensitivity of the reaction, the amount of linear DNA copy in each tube was determined and serial dilutions were prepared. Finally, LAMP reaction was performed on them. The obtained results showed that the threshold of the LAMP reaction was 10 copies of the target gene in each positive tube (Figure 3A).

**Figure 3.**
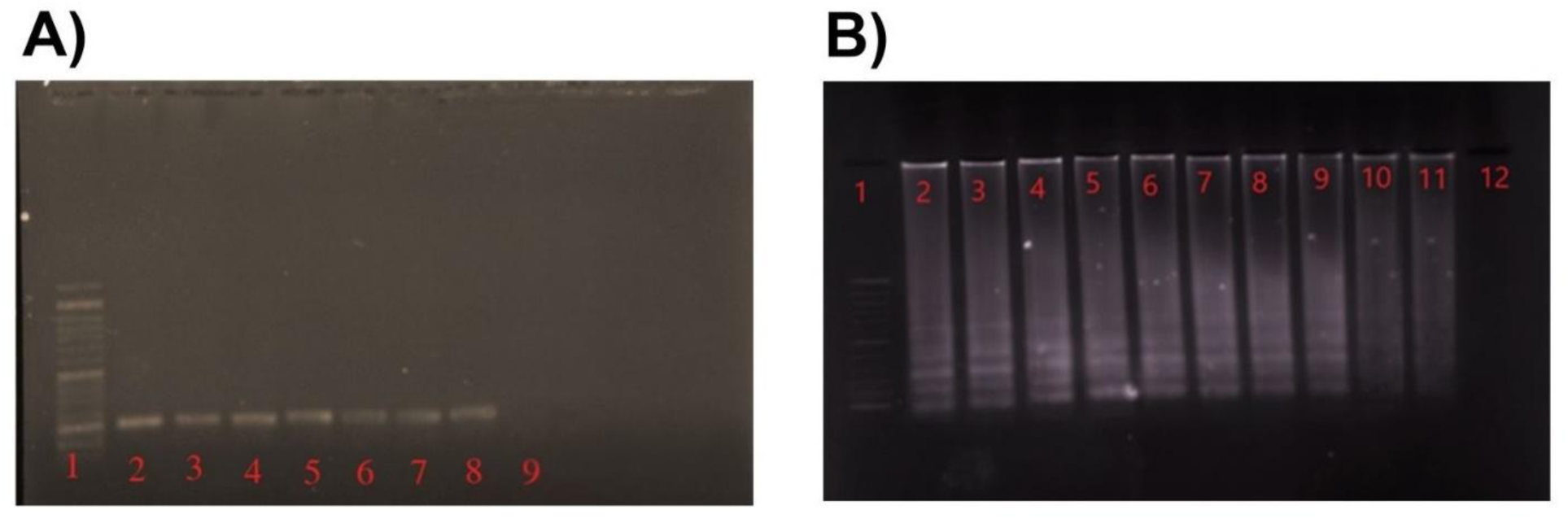
The sensitivity of *C. auris* detection via LAMP and PCR methods. The shown products are the 1) Ladder 50-kbp, 2-5) logarithmic dilutions of the linearized positive control from 10^2^ to 10^6^ of *C. auris*, 6) the dilution of 500 copies of *C. auris*, 7) the dilution of 250 copies of *C. auris*, 8) the dilution of 100 copies of *C. auris*, 9) the dilution of 10 copies of *C. auris*, 10) the dilution of 5 copies of *C. auris*, 11) the dilution of 1 copies of *C. auris*, and 12) negative control (without DNA sample) samples.

To check the sensitivity of the PCR method, the same dilutions of the LAMP technique were used. The results showed that the threshold of PCR detection was 100 copies/tube (Figure 3B).

### 3.4. Confirmation of LAMP test

#### Turbidity

The LAMP reaction was performed for 60 minutes at 65 °C in a heat block. As soon as the LAMP reaction was finished, the tubes were removed. The turbidity resulting from the formation of magnesium pyrophosphate deposit indicates a positive reaction that was observed with the eye in the positive tube, thus eliminating the need to perform gel electrophoresis (Figure 4A).

**Figure 4.**
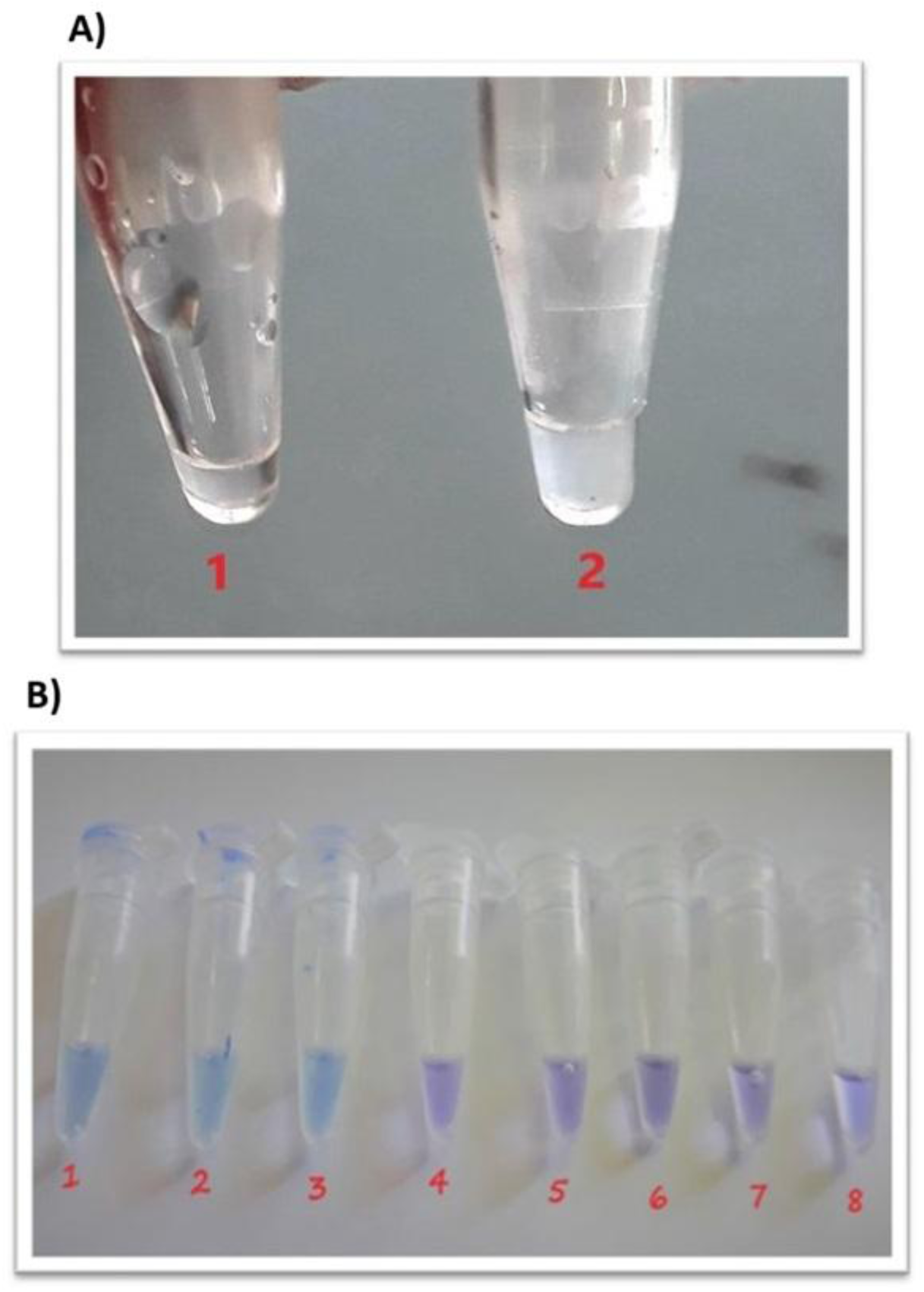
Turbidity and HNB tests after LAMP reaction. (A) The turbidity test was conducted and the results were shown in tube 1 as negative control and tube 2 as positive control (containing ITS genome). (B) The HNB test was conducted and the results were shown in 1) *C. auris* UMS44, 2) *C. auris* UMS48, 3*) C. auris* UMS51, 4) *C. albicans*, 5) *C. glabrata*, 6) *C. krusei*, 7) *C. parapsilosis*, and 8) *C. tropicalis.* HNB: hydroxynaphthol blue, LAMP: Loop-Mediated Isothermal Amplification.

#### HNB

The color of HNB is purple in the negative reaction tube, which changes colour to light blue in the positive reaction tube. The results are shown in Figure 4B.

### 3.5. DNA sequencing and nBLAST results

The results of sequencing determined a region of the gene with a length of 180 bp (Table 2) (Figure 5).

**Figure 5.**
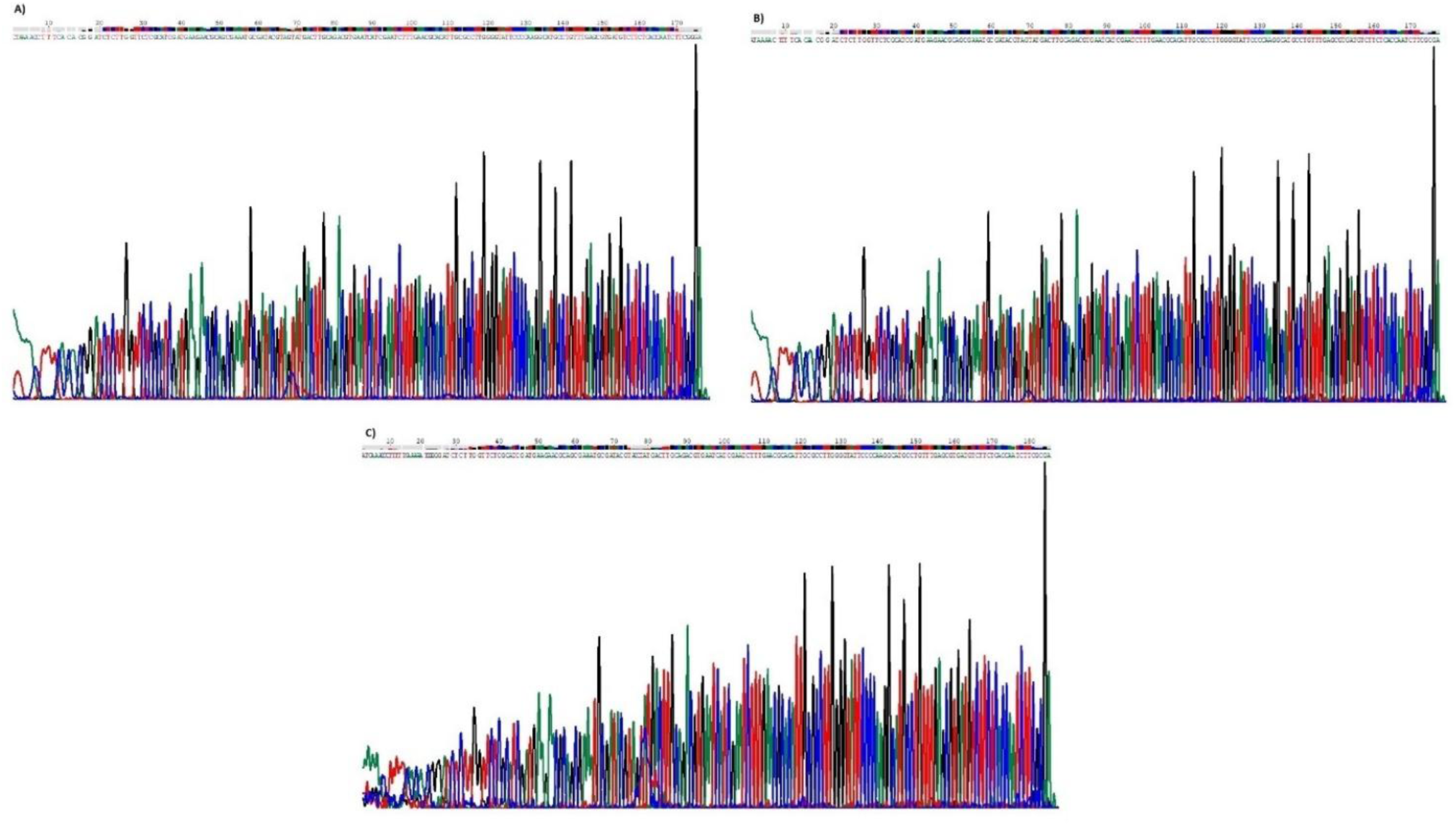
The chromatogram results. The chromatogram results of (A) *C. auris* UMS44, (B) *C. auris* UMS48, and *C. auris* UMS51 isolates were shown that were obtained via Choromas software.

**Table 2.**
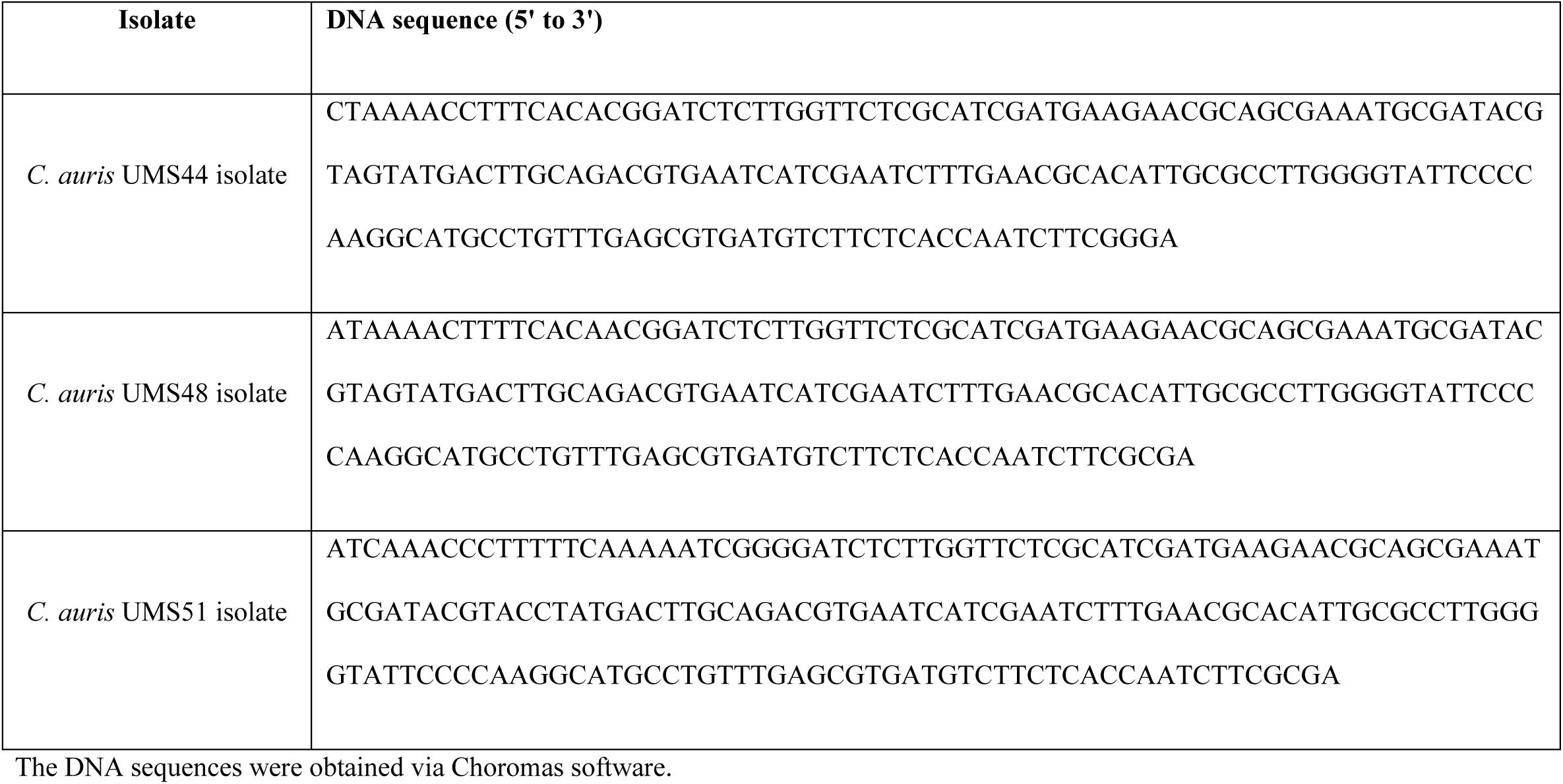
DNA sequences of PCR products.

By considering the percentage of overlap of the desired sequence (Query coverage) and the percentage of maximum similarity of their sequence (Max identity), the results obtained from the graphic nBLAST b showed that the sequences in the NCBI database were completely similar to the desired sequence. Therefore, with the results obtained from nBLAST, the studied isolates belonged to *C. auris* species. All three studied sequences showed a significant alignment for UMS44, UMS48, and UMS51 isolates, indicating a homology relationship with high certainty (Table 3).

**Table 3.**
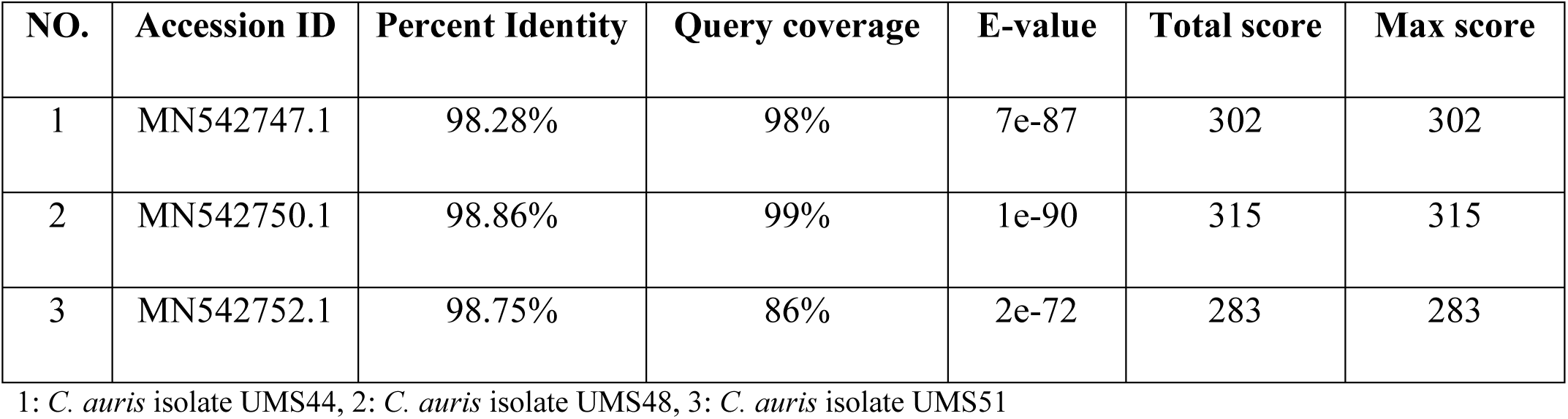
nBLAST results of studied positive samples.

| **NO.** | **Accession ID** | **Percent Identity** | **Query coverage** | **E-value** | **Total score** | **Max score** |
| --- | --- | --- | --- | --- | --- | --- |
| 1 | MN542747.1 | 98.28% | 98% | 7e-87 | 302 | 302 |
| 2 | MN542750.1 | 98.86% | 99% | 1e-90 | 315 | 315 |
| 3 | MN542752.1 | 98.75% | 86% | 2e-72 | 283 | 283 |

1: *C. auris* isolate UMS44, 2: *C. auris* isolate UMS48, 3: *C. auris* isolate UMS51

### 3.6. Phylogenic tree of Candida isolates

In this study, evolutionary history was evaluated using Mega6 software and cladogram drawing using Neighbor-Joining method, and evolutionary distances were calculated using Maximum Composite Likelihood method and bootstrap value of 1000.

Figure 6 shows the phylogeny relationship of the mentioned isolates and the isolates belonging to the reference *C. auris* available in GenBank. The results of the phylogeny tree show that the samples are placed in a clade with the support of 99, 92 and 72% bootstrap and with a small distance to each other, which indicates the monophyletic relationship between the studied isolates of *C. auris*. The ITS gene of *C. albicans*, *C. glabrata*, *C. krusei*, *C. parapsilosis*, and *C. tropicalis*, which was used as an outgroup, was placed in a separate branch from the others and rooted the tree.

**Figure 6.**
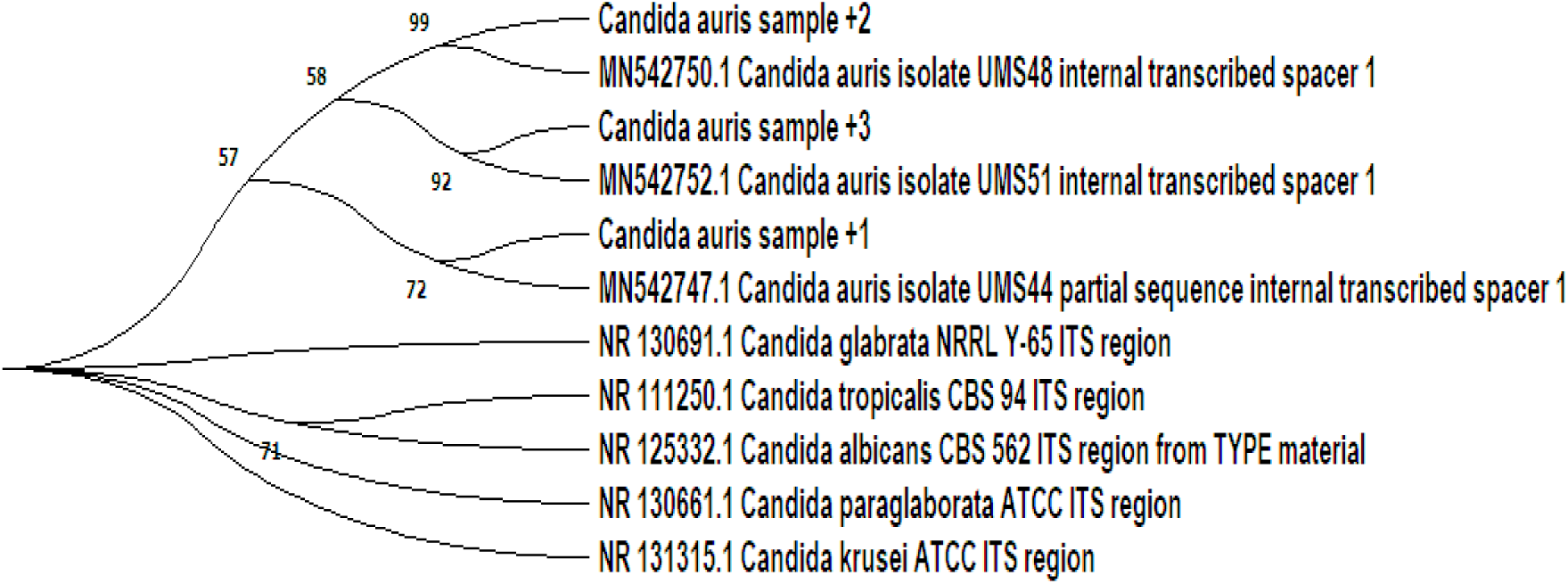
The phylogenic tree of Candida isolates.

## 4. Discussion

Quick and accurate diagnosis of *C. auris* along with evaluation of drug resistance are considered essential for disease management, infection prevention, and health control. The gold standard for detecting *C. auris* is PCR. Although the low quality and contamination in the DNA samples could decrease specificity and cause false positives in PCR, the LAMP reaction is not so sensitive to the quality and concentration of template DNA [10]. For the first time, Notomi et al developed a new method called LAMP that could selectively amplify DNA templates more faster than PCR [11]. LAMP is a new method to amplify the specific DNA sequence and it has successfully identified many fungal pathogens such as *Paracoccidioides brasiliensis*, *Fonsecaea species*, *Cladophialophora*, *C. albicans*, *C. glabrata*, *C. krusei*, and *C. lusitaniae* with 100% specificity and no cross-reaction with other closely related species [12]. In a recent metanalysis study by Bumbrah et al, it is illustrated that LAMP as a rapid, cheap, and repetitive method is a good alternative for human fungal detection which are not drug resistant [13].

In this project, we evaluated the isothermal LAMP method as an alternative method to the PCR method. The obtained results from this study demonstrated that LAMP can detect low copies of DNA in *C. auris* samples with specific primers that take less time than PCR. Moreover, it has high sensitivity and specificity in comparison to other species in the *Candida* family. The optimized temperature in this study was 65 °C which was consistent with the results of the research by Inamoto et al that used LAMP for HSV DNA detection from 18 samples of patients with gingivostomatitis/vesicular skin eruptions [14]. In a study by Poon et al that detected *Plasmodium falciparum* from blood samples of patients with Malaria by LAMP method, it is approved that there is no need to purify DNA samples before efficient DNA amplification in the LAMP method [15]. Accordingly, for obtaining sensitive results in PCR, it was shown that extraction and purification of DNA samples is necessary takes more time and money and it cannot be performed in all conditions [16]. In contrast, LAMP target gene detection from culture medium samples can be done without DNA extraction. It is revealed that LAMP has a higher tolerance to the culture medium and various biological fluids (plasma, serum, saline, urea, vitreous, etc.) compared to PCR [16]. Suzuki et al tested DNA samples of CMV infection in pediatric hematopoietic stem cell transplant recipients by LAMP and suggested 63-65 °C as the best temperature range for LAMP in CMV virus detection [17]. According to a study by Yeh et al that detected 18 gene of *Edwardsiella ictaluri* in the brains of channel catfish by LAMP, the minimum time for the LAMP reaction was predicted 60 minutes to obtain a valid product [18]. However, in a study by Nagamine et al, LAMP was introduced as a high-yielded technique and it was demonstrated that cyclic initiators could reduce the time to 30 minutes of detection by turbidity measurement [19]. Another study by Hayashi et al also showed that using cyclic initiators at 65°C can accelerate the LAMP reaction [20]. Goodarzi and his colleagues compared the LAMP method and PCR to identify *C. albicans* in pharyngeal secretions of suspected tuberculosis patients [21]. The PCR method detected at least 100 copies of DNA samples and the LAMP method detected at least 10 copies in a 25 µL reaction, similar to our findings. In another study by Yamamoto et al, it is indicated that LAMP could detect *C. auris* strains from other strains with 100% specificity, and the threshold DNA concentration was 20 copies per reaction [22].

## 5. Conclusion

Based on our results, the adapted LAMP method could identify all isolated *C. auris* from other related species with 100% specificity and no cross-reactions were observed during the experiments. Moreover, because of using turbidity and HNB measurement instead of electrophoresis, the final detection time of the LAMP method was much shorter than PCR. Because of the simple process of LAMP, this method could be performed by even non-specialists and lead to early diagnosis and effective treatment of related hospital infections caused by *C. auris,* and the final cost of LAMP is much lower than PCR. The LAMP technique had high sensitivity compared to PCR and could even detect 10 copies of the target DNA in a reaction in the early stages of an infection. The repeatability of the LAMP method was also confirmed by performing on different days. One of the main challenges in detecting *C. auris* is accurate differentiation of its genome from the closely related *Candida* species, and our study holds primes for accurate detection of *C. auris* with great accuracy and a sensitivity surpassing PCR. This study can be used as basis for developing POC molecular assay for *C. auris* via studying clinical samples which were not feasible in the current study.

## Declarations Ethical approval

This project was approved by the committee for ethics in biomedical research of Tabriz University of Medical Sciences (Ethical code: IR.TBZMED.REC.1396.1200) and it was carried out according to the international

## CRediT authorship contribution statement

Conceptualization, MPM; Methodology, MPM, LM; Software, LM; Formal analysis, LM; Funding acquisition, MPM; Investigation, MPM, LM; Resources, LM; Data curation, LM; Supervision, MPM; Project administration, MPM, LM; Visualization, LM; Validation, LM; Writing—Original Draft, LM; Writing—Review & Editing, LM.

## Data Availability

All data produced in the present study are available upon reasonable request to the authors

## Acknowledgments

None.

## Conflict of Interest

The authors declare that they have no competing interests.

## Data Availability

Data will be made available upon reasonable request.

## Funding supports

This study was financially supported by Tabriz University of Medical Sciences (#Grant No.: 59182).

## Notes

### Competing Interest Statement

The authors have declared no competing interest.

## References

[1] M. A. P. Medeiros, A. P. V. Melo, A. O. Bento, L. Souza, F. A. B. Neto, J. B. Garcia, D. L. Zuza-Alves, E. C. Francisco, A. S. A. Melo, G. M. Chaves Epidemiology and prognostic factors of nosocomial candidemia in Northeast Brazil: A six-year retrospective study. PLoS One, 14 (8) (2019), e0221033. 10.1371/journal.pone.0221033

[2] J. de Cássia Orlandi Sardi, D. R. Silva, M. J. Soares Mendes-Giannini, P. L. Rosalen *Candida auris*: Epidemiology, risk factors, virulence, resistance, and therapeutic options. Microb Pathog, 125 (2018), 116–121. 10.1016/j.micpath.2018.09.014

[3] F. Di Serio, G. Antonelli, P. Trerotoli, M. Tampoia, A. Matarrese, N. Pansini Appropriateness of point-of-care testing (POCT) in an emergency department. Clin Chim Acta, 333 (2) (2003), 185–189. 10.1016/s0009-8981(03)00184-0

[4] P. B. Luppa, C. Müller, A. Schlichtiger, H. Schlebusch Point-of-care testing (POCT): Current techniques and future perspectives. Trends Analyt Chem, 30 (6) (2011), 887–898. 10.1016/j.trac.2011.01.019

[5] P. Craw, W. Balachandran Isothermal nucleic acid amplification technologies for point-of-care diagnostics: a critical review. Lab Chip, 12 (14) (2012), 2469–2486. 10.1039/c2lc40100b

[6] S. K. Vashist, P. B. Luppa, L. Y. Yeo, A. Ozcan, J. H. T. Luong Emerging Technologies for Next-Generation Point-of-Care Testing. Trends Biotechnol, 33 (11) (2015), 692–705. 10.1016/j.tibtech.2015.09.001

[7] R. Hans, N. Marwaha Nucleic acid testing-benefits and constraints. Asian J Transfus Sci, 8 (1) (2014), 2–3. 10.4103/0973-6247.126679

[8] P. A. Gaudio, U. Gopinathan, V. Sangwan, T. E. Hughes Polymerase chain reaction based detection of fungi in infected corneas. Br J Ophthalmol, 86 (7) (2002), 755–760. 10.1136/bjo.86.7.755

[9] S. Petralia, S. Conoci PCR Technologies for Point of Care Testing: Progress and Perspectives. ACS Sens, 2 (7) (2017), 876–891. 10.1021/acssensors.7b00299

[10] N. Kuboki, N. Inoue, T. Sakurai, F. Di Cello, D. J. Grab, H. Suzuki, C. Sugimoto, I. Igarashi Loop-mediated isothermal amplification for detection of African trypanosomes. J Clin Microbiol, 41 (12) (2003), 5517–5524. 10.1128/jcm.41.12.5517-5524.2003

[11] T. Notomi, H. Okayama, H. Masubuchi, T. Yonekawa, K. Watanabe, N. Amino, T. Hase Loop-mediated isothermal amplification of DNA. Nucleic Acids Res, 28 (12) (2000), E63. 10.1093/nar/28.12.e63

[12] D. Biswal Advances in Loop-Mediated Isothermal Amplification (LAMP) Technology and Its Necessity to Detect Helminth Infections: An Overview. (2017). 10.21767/2472-1646.100015

[13] G. S. Bumbrah, S. Jain, S. Singh, Z. Fatima, S. Hameed Diagnostic Efficacy of LAMP Assay for Human Fungal Pathogens: a Systematic Review and Meta-analysis. Curr Fungal Infect Rep, (2023), 1–11. 10.1007/s12281-023-00466-0

[14] Y. Enomoto, T. Yoshikawa, M. Ihira, S. Akimoto, F. Miyake, C. Usui, S. Suga, K. Suzuki, T. Kawana, Y. Nishiyama, Y. Asano Rapid diagnosis of herpes simplex virus infection by a loop-mediated isothermal amplification method. J Clin Microbiol, 43 (2) (2005), 951–955. 10.1128/jcm.43.2.951-955.2005

[15] L. L. Poon, B. W. Wong, E. H. Ma, K. H. Chan, L. M. Chow, W. Abeyewickreme, N. Tangpukdee, K. Y. Yuen, Y. Guan, S. Looareesuwan, J. S. Peiris Sensitive and inexpensive molecular test for falciparum malaria: detecting Plasmodium falciparum DNA directly from heat-treated blood by loop-mediated isothermal amplification. Clin Chem, 52 (2) (2006), 303–306. 10.1373/clinchem.2005.057901

[16] H. Kaneko, T. Kawana, E. Fukushima, T. Suzutani Tolerance of loop-mediated isothermal amplification to a culture medium and biological substances. J Biochem Biophys Methods, 70 (3) (2007), 499–501. 10.1016/j.jbbm.2006.08.008

[17] R. Suzuki, T. Yoshikawa, M. Ihira, Y. Enomoto, S. Inagaki, K. Matsumoto, K. Kato, K. Kudo, S. Kojima, Y. Asano Development of the loop-mediated isothermal amplification method for rapid detection of cytomegalovirus DNA. J Virol Methods, 132 (1-2) (2006), 216–221. 10.1016/j.jviromet.2005.09.008

[18] H. Y. Yeh, C. A. Shoemaker, P. H. Klesius Evaluation of a loop-mediated isothermal amplification method for rapid detection of channel catfish Ictalurus punctatus important bacterial pathogen Edwardsiella ictaluri. J Microbiol Methods, 63 (1) (2005), 36–44. 10.1016/j.mimet.2005.02.015

[19] K. Nagamine, T. Hase, T. Notomi Accelerated reaction by loop-mediated isothermal amplification using loop primers. Mol Cell Probes, 16 (3) (2002), 223–229. 10.1006/mcpr.2002.0415

[20] N. Hayashi, R. Arai, S. Tada, H. Taguchi, Y. Ogawa Detection and identification of Brettanomyces/Dekkera sp. yeasts with a loop-mediated isothermal amplification method. Food Microbiol, 24 (7-8) (2007), 778–785. 10.1016/j.fm.2007.01.007

[21] M. Goodarzi, M. H. Shahhosseiny, M. Bayat, S. J. Hashemi, M. Ghahri Comparison between Molecular Methods (PCR vs LAMP) to Detect Candida albicans in Bronchoalveolar Lavage Samples of Suspected Tuberculosis Patients. Microbiology Research, 8 (2) (2017), 7306.

[22] M. Yamamoto, M. M. Alshahni, T. Tamura, K. Satoh, S. Iguchi, K. Kikuchi, M. Mimaki, K. Makimura Rapid Detection of *Candida auris* Based on Loop-Mediated Isothermal Amplification (LAMP). J Clin Microbiol, 56 (9) (2018). 10.1128/jcm.00591-18

